# Environmental pollutants and essential elements as regulators of miR-30b, miR-223 and Let-7a microRNAs expression in maternal sera from the NEHO cohort

**DOI:** 10.1101/2022.01.31.22270144

**Authors:** Valeria Longo, Gaspare Drago, Alessandra Longo, Silvia Ruggieri, Mario Sprovieri, Fabio Cibella, Paolo Colombo

## Abstract

**Background:** Birth cohorts of women living in highly industrialized areas provide an ideal setting for studying the correlation between environmental exposure and health.

**Objectives:** To evaluate serum microRNA expression in response to environmental contaminants in 68 healthy pregnant women from the NEHO birth cohort.

**Methods:** Serum contaminants were determined by GC-MS/MS and ICP-MS/MS. Serum microRNA expression was determined by qPCR. Associations between miRNA ΔCTs and single serum element levels were tested by linear regression models, while whole mixture effect was studied by WQS regression analysis.

**Results:** In separate regression models, a positive association was found between miR-223 ΔCT and concentrations of Se and Zn, while a negative association between miR-30b and Hg levels was observed. Similar regression models were also conducted using tertiles of each chemical as independent variables. In this setting, the Let-7a ΔCT was decreased in the comparison between medium and low tertiles of Se. The highest tertiles of Zn and p,p’-DDE showed a significant association with increasing levels of miR-223 ΔCT with respect to the lower tertile; moreover, miR-30b ΔCT was reduced in the comparison between high and low tertiles of Hg. The observed data were confirmed by fold-change analysis. A WQS analysis built on tertiles of contaminant distribution revealed a significant mixture effect on the expression of the analyzed miRNAs. The inverse association between Let-7a ΔCT and the WQS index was dominated by Zn, Se, Cu, Hg and HCB. Moreover, miR-223 ΔCT was positively associated with the WQS index, where p,p’-DDE, Zn and Se showed the greatest contributions to the association. Conversely, the reduction of miR-30b ΔCT was mainly driven by Hg and Se.

**Discussion:** This study allowed us to characterize the role of prolonged exposure to environmental contaminants influencing the expression of circulating miRNAs in the serum of women in the last trimester of pregnancy.

## INTRODUCTION

The exponential growth of the world economy and, with that, exploitation of resources and industrial production caused an unprecedented transfer of contaminants in the environment with multiple impacts on health. These chemicals can enter the human body through various routes: ingestion of chemicals via the consumption of contaminated food or water, inhalation of airborne pollutants through the respiratory system as well as dermal absorption through the skin. This presents a highly problematic issue in everyday life and, particularly, in pregnancy due to potential fetal exposure during a very vulnerable phase of development. A paradigmatic example is the adverse effects on the child from high maternal prenatal mercury exposure, as reported in the Minamata accident, with an overwhelming incidence of symptomatic mothers giving birth to children with serious neurologic diseases (Ekino et al. 2007). Life *in utero* is considered a particularly sensitive period during which maternal exposure to unfavorable conditions may not only influence fetal development and induce adverse pregnancy outcomes but also have long-term effects influencing offspring susceptibility to diseases later in adulthood. This causality concept is known as the Developmental Origins of Health and Disease (DOHaD) or Barker hypothesis (Barker 1993). Various studies have already described associations between prenatal toxicant exposures and impaired birth and childhood outcomes (Barouki et al. 2012; Deng et al. 2016; Lamichhane et al. 2015; Rahman et al. 2015; Stolevik et al. 2013) showing the extraordinary power of prospective pregnancy or birth cohort studies to examine the potential health effects of developmental exposure to chemicals. In this respect, birth cohort studies provide the most suitable design for assessing the association of early-life adversities occurring at critical developmental windows with their possible long-lasting effects on postnatal health and well-being.

Recently, the International Centre of Advanced Study in Environment, Ecosystem and Human Health (CISAS) has developed a project (http://www.cisas.cnr.it/) aimed at investigating environmental pollution and its connection with the ecosystem and human health in highly contaminated areas of southern Italy monitoring pollutant concentrations in human tissues and also performing *in vitro/ex vivo* studies (Bocca et al. 2020; Drago et al. 2020; Longo et al. 2019; Longo et al. 2021; Montalbano et al. 2020; Traina et al. 2021).

Augusta Bay is a ∼25km^2^ marine coastal area, located in southern Italy (Figure 1) and hosting, since 1950s, one of the largest petrochemical plant in Europe, with associated oil refineries and a chloralkali plant. Decades of uncontrolled industrial discharges led to a major historical contamination of sediments with mixtures of trace metals (Hg, Methyl-Hg, Pb, Cu, Zn, Cd, etc.) and organic compounds (mainly, aliphatic hydrocarbons, PCBs, PAHs and HCB) (Bellucci et al. 2012; Salvagio Manta et al. 2016; Sprovieri et al. 2011). Also, a relevant number of pharmaceutical products were detected at relatively high toxic levels in recent sediments of the bay while a number of classes of ionic surfactants (alkylsulfonic acids, alkylbenzensulfonic acids, dodecyl-and tetradecyl-polyethoxy sulfates), non-ionic surfactants (polyethoxylated alcohols) and personal care components (cocamidopropylbetaine, dodecyldimethylamine oxide, cocamide- and similar compounds) were measured in seawater (Feo et al. 2020). Significant mercury evasion processes to the atmosphere were quantitatively estimated (Bagnato et al. 2013; Sprovieri et al. 2011). Bioaccumulation of contaminants in marine edible organisms were reported by Traina and coworkers (Traina et al. 2021) while Di Bella and coworkers (Di Bella et al. 2020) documented effects of multiple pollution on commercial milk and meat samples collected from local markets in surrounding area. This highly heterogeneous system of pollution sources and pathways characterizing the Augusta area offers a multi-exposure setting with a highly complex (and only partially classified and quantified) mixture of pollutants that directly affect the residing population with major effects of health implications (Zona et al. 2019). Nevertheless, establishing pathophysiological causal links between environmental pollutants and health consequences remains a priority challenge due to the multi-factorial dynamics of exposures (Mudu et al. 2014). To define the totality of environmental exposures, the concept of exposome has been developed on the attempt to identify chemical pattern of exposure relevant for adverse health effects, thus complementing the contribution of life style factors and genomic susceptibility to human disease development (Wild 2005).

**Figure 1.**
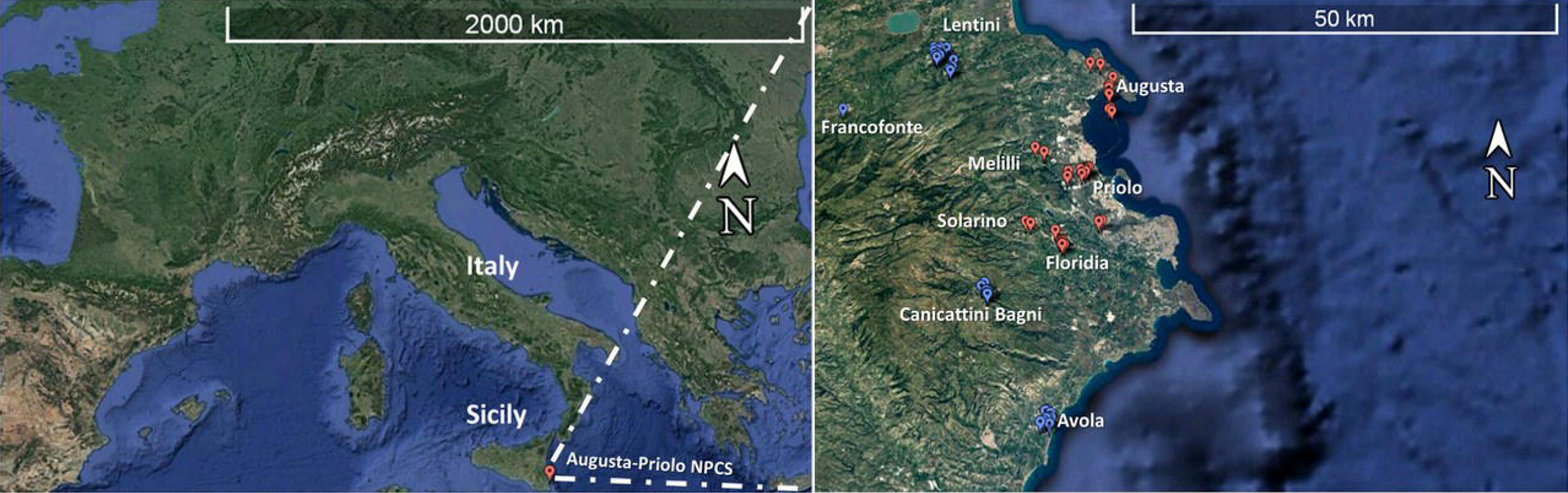
Geographical localization of the study area. Left panel shows the geographical localization of the Priolo-Augusta study area. The Right panel shows the localization of the dwellings of the mothers in the area. Mothers residing in National Priority Contaminated Sites are in red, mothers residing in the Local Reference Areas are in blue.

Biomonitoring of organic and inorganic substances in the blood of people is an ideal approach to investigating causal relationships between environmental exposure and human health. Moreover, biomonitoring allows direct assessment of the distribution of risk factors in the population, integrating the individual variability of exposures, as well as the different capacity to metabolize and transform the chemicals present in the body (Louro et al. 2019). Only a small portion of risk for chronic diseases can be explained by genetic factors, while a large portion of the remaining risk can be attributed to the impact of contaminants found in the blood (Rappaport 2016). Human blood is a commonly used sample matrix in clinical as well as epidemiological studies. A large set of chemicals from food, drugs, household chemicals, and environmental pollutants can enter the bloodstream and affect cell viability. The concept of ‘blood exposome’ includes investigation of association models on potentially all the chemical contaminants measured in human blood (Rappaport et al. 2014) and interpreting this distribution in terms of phenotypes and physiology impact and to provide robust estimates of risk for acute and chronic diseases (Dennis et al. 2017).

Mammalian genes are susceptible to changes depending on their environment, influencing disease development and promoting perdurable epigenetic changes (Junien et al. 2016). Such epigenetic modification includes histone modification, DNA methylation and microRNA (miRNA) gene silencing. All of these are closely linked to each other and influence protein synthesis patterns whose perturbation may lead to dysfunction. Currently, many studies suggested that epigenetic modification can play an important role in the pathogenesis of some multifactorial diseases (Apicella et al. 2019; Arck and Hecher 2013; Boosani and Agrawal 2016).

MicroRNAs are highly conserved small single-stranded non-coding RNA (18 ∼ 25 nucleotides) considered a major category among the noncoding RNA fraction that regulates gene expression at the post-transcriptional level (Miguel et al. 2020). Different miRNAs are associated with several metabolic pathways, including lipid metabolism, glucose metabolism, food intake, body weight homeostasis, inflammation, oxidative stress, expression of several cytokines, and angiogenesis in obesity. Circulating microRNAs may reflect these alterations in tissue expression and intracellular signaling, supporting their utility as novel and minimally invasive biomarkers of environmental exposure to pollutants (Wu et al. 2021).

In this study, a random sample of 68 pregnant women, living in the Augusta area, was enrolled from the NEHO cohort (Ruggieri et al. 2019). A multipollutant analysis was performed to explore the correlation between the expression levels of miR-30b, miR-223 and Let-7a and the presence of a suite of inorganic and organic elements in the sera of such women. The aim of the study was to evaluate perturbations in the expression of relevant circulating immune-related miRNAs in women in the last trimester of pregnancy in response to long-lasting chronic environmental exposure.

## MATERIAL AND METHODS

### Study population

The present study included 68 women followed up in the larger context of the NEHO birth cohort. The studied individuals reside in the Augusta-Priolo area officially included in the National Remediation Plan by the Italian Ministry of Environment (NPCS) and in a surrounding area (Local Reference Area [LRA]) reported as a ‘local control’ (Carere et al. 2016; Zona et al. 2019). The NEHO cohort methods are described in detail elsewhere (Ruggieri et al. 2021). Briefly, between January 2018 and January 2020, the NEHO cohort enrolled 561 pregnant women residing, for at least the last year before enrollment, in the NPCS and LRA, with comparable socio-demographic characteristics. Detailed information was collected from the mothers using web-based questionnaires at enrollment as well as at 6, 12, and 24 months after delivery. Moreover, general and clinical details about the type of delivery and birth outcome (i.e. weight, length, head circumference) were collected by healthcare personnel at delivery. Maternal blood samples were gathered at enrollment. All mothers included in this research do not evidence i) any specific chronic diseases (such as diabetes, hypertension, etc.) and ii) any critical complications during pregnancy diagnosed previous to signing informed consent.

### Blood sampling and handling

After separation of the serum by centrifugation, the samples were temporarily stored at -20°C in each maternal unit and then transported on dry ice to the NEHO biobank for long-term storage at - 80°C.

The study was approved by the relevant Ethics Committee (Comitato Etico Catania 2, 11 July 2017, n. 38/2017/CECT2), and strictly followed the Declaration of Helsinki. All the adopted procedures were compliant with the General Data Protection Regulation (UE 2016/679) and Italian laws concerning data protection. To this aim, all questionnaire data and biological samples were pseudonymized using ID tracking numbers.

### Analytical methods for trace metals and organic compounds

The analyses of Persistent Organic Pollutant (POP) were performed at the National Institute for Health and Welfare, Chemical Exposure Unit, Kuopio, Finland, with an Agilent 7000B gas chromatograph triple quadrupole mass spectrometer (GC-MS/MS). Ethanol and ^13^C-labeled internal standards were added to samples. Dichloromethane-hexane was added for extraction, followed by the addition of activated silica gel to bind the sample water and ethanol. The dichloromethane-hexane layer was poured into a solid phase extraction cartridge (SPE cartridge) containing 44% sulfuric acid silica, 10% silver nitrate impregnated silica, and a mixture of sodium sulfate and silica. The lower semi-solid layer was extracted again with dichloromethane-hexane that was also poured into an SPE-cartridge. Elution of the SPE-cartridge was continued with dichloromethane-hexane and the eluate was concentrated for GC-MS/MS. The quantification was performed by multiple reaction monitoring using an Agilent 7890A gas chromatograph/Agilent 7010 triple quadrupole mass spectrometer with DB-5MS UI column (J&W Scientific, 20 m, lD 0.18 mm, 0.18 μm). Reference material for human serum determination of organic contaminants (SMR 1589a) from the National Institute of Standards and Technology (Gaithersburg, MD, USA) was analysed to estimate the accuracy: recoveries for each analyte of PCBs, p,p’-DDE and HCB ranged between 94% and 104%. Analytical precision was routinely better than 3% RSD%.

Analyses of trace element were performed in the micropollutant unit laboratory of LERES (Laboratoire d’Etude et de Recherche en Environnement et Santé) at the French School of Public Health - EHESP (Rennes, France) following the procedures reported by (Davies et al. 2021). The 68 serum samples were analyzed by ICP-MS/MS after a mineralization step. Briefly, a volume of serum was mineralized by adding nitric acid and heated with a heating block (Hotblock Pro, model SC-189, Environmental Express) at 83°C for 4 hours. The 1:20-diluted sample was then analyzed by a plasma inductive system coupled with tandem mass spectrometry detection (ICP-MSMS 8800, Agilent Technologie). The use of the tandem mass spectrometer and oxygen mode allowed interference to be managed, in particular for the determination of manganese, selenium and arsenic. Matrix effects correction has been guaranteed with the use of internal standards (Sc, Ge, ^77^Se, Rh, Re and Ir). This made possible to check the absence of drift (signal variation less than 25%). For ^77^Se an internal standard was added to the mixture to allow quantification of Se by isotopic dilution to resolve persistent matrix effects. Certified or internal control materials (measured additions) of blood and serum were added to the series.

### Serum miRNA extraction and expression analysis

Total RNAs enriched with miRNAs were extracted from 200 μl of maternal serum samples according to the miRNeasy Serum/Plasma Kit manufacturer’s protocol (Qiagen, Milan, Italy). The *Caenorhabditis elegans* miR-39 exogenous control (Qiagen, Milan, Italy) was added during the extraction procedure in all samples (3.5 μl/sample at a concentration of 1.6×10^8^ copies/μl). The total RNA and miRNA concentrations were evaluated by NanoDrop spectrophotometer analysis (Thermo Fisher Scientific, Monza, Italy). The templates (500 ng for each samples) were retro-transcribed using the miScript II RT Kit (Qiagen, Milan, Italy), and the resulting cDNAs were diluted up to 200 μl. Then, the real time PCR analyses (StepOnePlusTM Real Time PCR System, Applied Biosystems) were performed starting from 1 ng of cDNA/well using the SYBR® Green PCR miScript kit (Qiagen, Milan, Italy); each reaction was in a final volume of 25 μl/well. The real time PCR conditions were: 95°C for 15 minutes followed by 40 cycles of three step PCR; denaturation at 94°C for 15 seconds; annealing at 55°C for 30 seconds; extension at 70°C for 30 seconds. The expression levels for the selected miRNAs (miR-30b, miR-223 and Let-7a) were also reported as fold changes using the 2^-ΔΔCT^ method. MiRNA expression was considered changed between the two groups (control and treated cells, respectively) if it met the following criteria: fold change <0.5 (down-regulated), fold change >2 (up-regulated), quantification cycle (CT) value <35.

### Statistics

Continuous variables are reported as mean ± SD or median and interquartile range (IQR), for normally and non-normally distributed variables, respectively, while categorical variables are reported as numbers and percentages. For continuous variables, one-way ANOVA and the Mann-Whitney *U* test were used to test differences between the NPCS and LRA, when appropriate. A chi-squared test was used to study the association between area of residence (NPCS and LRA) and categorical variables.

The Spearman rank correlation test was used to check for possible pairwise correlations between each log_2_ transformed contaminant concentration, and the miRNA ΔCT. A multiple linear regression model was used to test the possible association between log_2_ concentration of bio-monitored substances and miRNA ΔCT. Each model was corrected by maternal age and Body Mass Index (BMI).

To account for the collinearity among the available contaminants, Weighted Quantile Sum (WQS) regression analysis was conducted to derive the mixture indices associated with miRNA ΔCT values using the “gWQS: generalized Weighted Quantile Sum regression” package in R. Two separate WQS indices were derived: an index positively associated with ΔCT of each considered miRNA, and an index negatively associated with ΔCT. A linear link function connecting outcome mean to a weighted sum of exposure tertiles was used.

Multiple linear regression models were created for each substance using the categorization into tertiles, using the lowest tertile as reference category. Also in this case, each model was adjusted for maternal age and BMI.

All tests were conducted at a nominal alpha error of 0.05, and no correction for multiple comparisons was performed. All the analyses were performed in R software, version 4.0.2

## RESULTS

### Characteristics of study population

In this study, 68 healthy pregnant women, enrolled in the NEHO birth cohort and residing in both the NPCS (n= 33) and LRA (n=35) of the Augusta-Priolo study site, were randomly selected. The geographical distribution of places of residence is reported in Figure 1. The analysis of demographic and gestational information (Table 1) shows that the two groups of mothers are homogeneous for age, BMI, weight gain, marital status, educational levels, gestational length (weeks), and birth type (Caesarean section or vaginal).

**Table 1.** Demographic and gestational characteristics of the study population.

|  | NPCS<br>n=33 | LRA<br>n=35 | p value* | NEHO<br>n=68 |
| --- | --- | --- | --- | --- |
| **Maternal characteristics** |  |  |  |  |
| **Age** (year, mean [±SD]) | 31.2 [±4.3] | 30.9 [±4.4] | 0.816 | 31.0 [±4.3] |
| **BMI** (kg/m<sup>2</sup>, mean [±SD]) | 23.7 [±5.7] | 23.3 [±5.7] | 0.902 | 25.5 [±5.7] |
| **Weight Gain** (kg, mean [±SD]) | 11.8 [±4.2] | 13.5 [±4.7] | 0.219 | 12.7 [±4.7] |
| **Marital status** (N [%]) |  |  | 0.875 |  |
| Married | 21 [63.6%] | 23 [65.7%] |  | 44 [64.7%] |
| Never Married/Separated | 12 [36.4%] | 12 [34.3%] |  | 24 [35.3%] |
| **Educational level** (N [%]) |  |  | 0.080 |  |
| Secondary school or lower qualification | 5 [15.2%] | 11 [31.4%] |  | 16 [23.5%] |
| High School | 22 [66.7%] | 14 [40.0%] |  | 36 [53.0%] |
| Degree or higher qualification | 6 [18.2%] | 10 [28.6%] |  | 16 [23.5%] |
| **Length of gestation** (weeks, mean [±SD]) | 39.2 [±1.3] | 39.1 [±1.2] | 0.346 | 39.1 [±1.2] |
| **Birth type** (N [%]) |  |  |  |  |
| Cesarean section | 14 [42.4%] | 11 [31.4%] |  | 25 [36.8%] |
| Vaginal | 19 [57.6%] | 24 [68.6%] |  | 43 [63.2%] |
| **Newborn Characteristics** |  |  |  |  |
| **Gender** (N [%]) |  |  | 0.839 |  |
| Female | 14 [42.4%] | 14 [40.0%] |  | 28 [41.2%] |
| Male | 19 [57.6%] | 21 [60.0%] |  | 40 [58.8%] |

\*p-Values comparing NPCSSs and LRAs based on Mann-Whitney U test for continuous variables and Chi-Square test for categorical variables.

### Evaluation of maternal serum contaminant levels

The dosage of a total of 10 POPs (HCB, p,p’-DDE, PCB118, PCB138, PCB153, PCB180), inorganic pollutants, and essential trace elements (Hg, Cu, Se, and Zn) was carried out on maternal serum samples (n=68) as above described. The concentrations of trace metals and POPs are reported in Table 2, with basic statistics. The Mann-Whitney *U* test highlighted a significant difference between mothers from the NPCS and LRA for Cu (p=0.008), Se (p=0.002), and Zn (p=0.003).

**Table 2.**
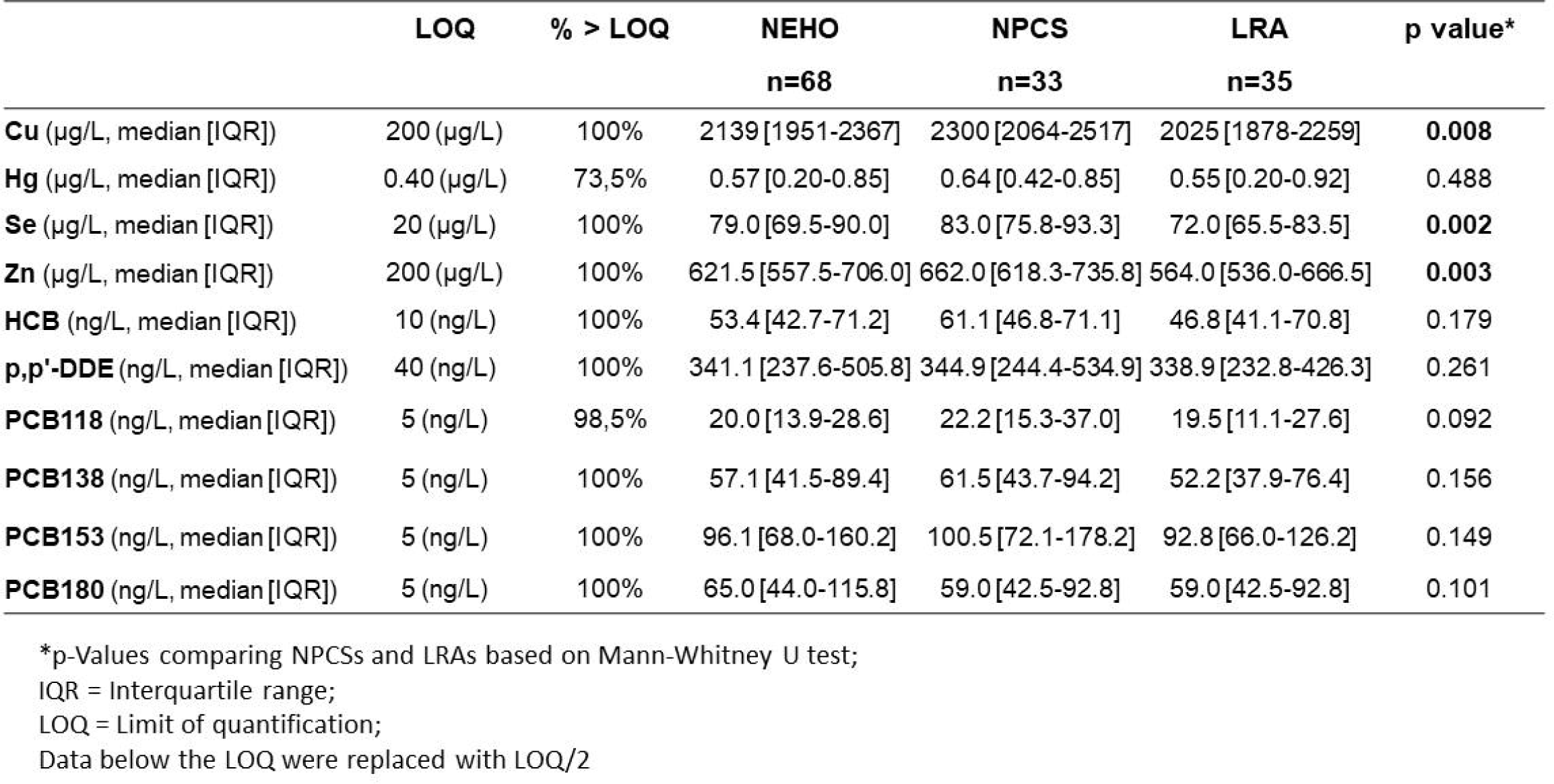
Serum maternal toxicant levels. Values are expressed as median and interquartile range.

### Analysis of miR-223, miR-30b and Let-7a in maternal serum

To evaluate possible epigenetic modifications induced by contaminants exposure, the maternal serum levels of relevant miRNAs were investigated. In particular, we analyzed the expression levels of miR-223, miR-30b, and Let-7a by real time PCR in the 68 serum samples. The Mann-Whitney *U* test (Supplementary Table 1) did not show significant differences in miRNA ΔCT values between the mothers from NPCS and LRA (miR-223 p=0.109; miR-30b p=0.729 and Let-7a p=0.470). These data are confirmed by the EDA plot displayed in Figure 1s, where the serum miRNA ΔCT are plotted according their geographical distributions, expression levels, and relative frequencies.

The dependence between multiple variables in the group of mothers was investigated by means of a Pearson’s correlation matrix between the miRNA expression levels and the concentration of maternal trace elements. The results showed a weakly negative and significant correlation between miR-30b and Hg (ρ=-0.28; p <0.0048). Similarly, miR-223 appears correlated to serum Se and Zn concentrations (ρ=0.28 and ρ=0.32, respectively). Moreover, among the evaluated environmental toxicants, PCBs showed a strong pairwise correlation among them (ranging from 0.60 to 0.98), also showing correlations to other POPs (HCB and p,p’-DDE), with correlation coefficients ranging from 0.69 to 0.75, respectively (see Figure 2 for details).

**Figure 2.**
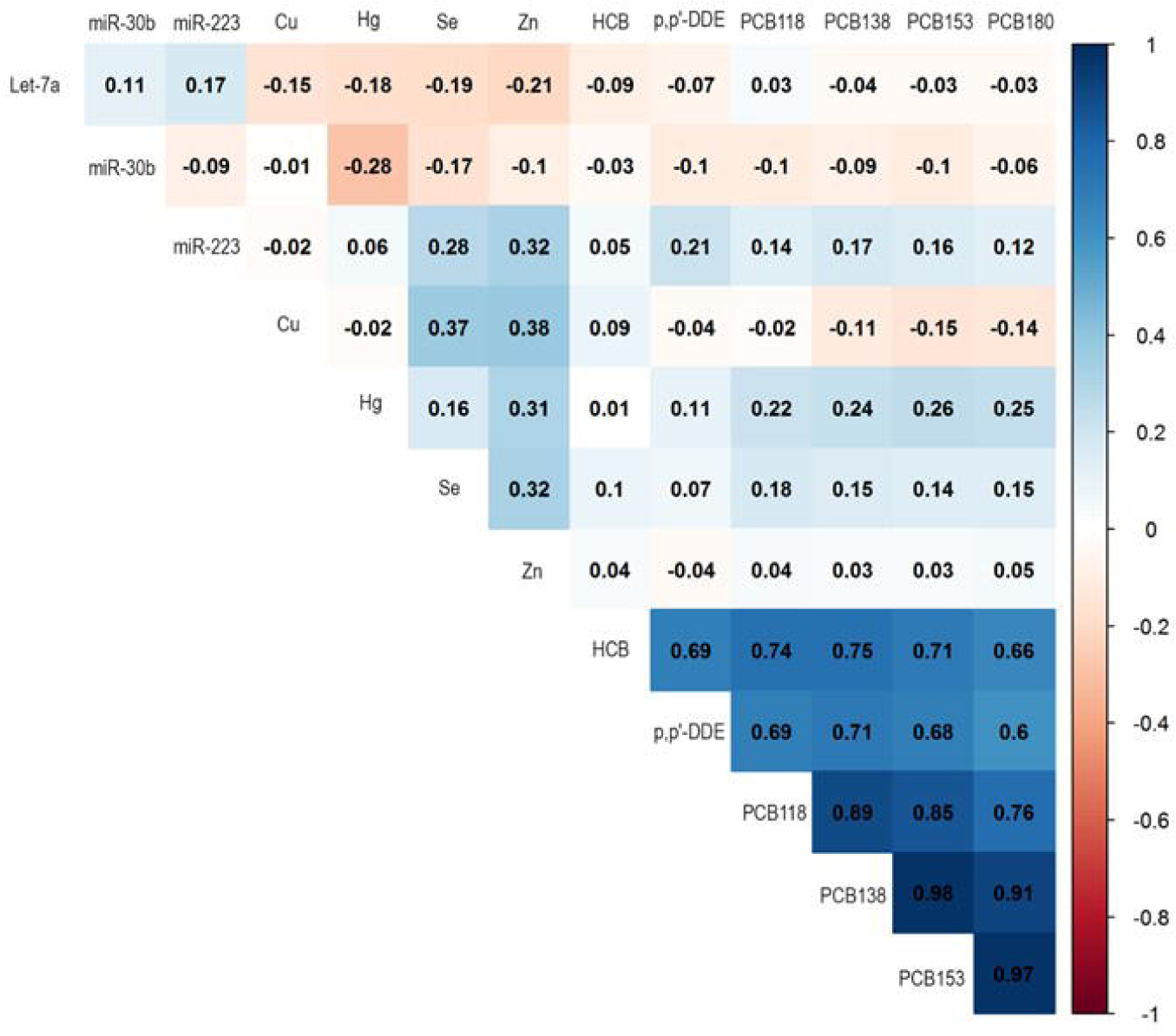
Pearson’s correlation matrix. miRNA ΔCTs and maternal serum levels of environmental toxicants and essential trace elements were analyzed in a correlation matrix. Correlations with a p-value > 0.05 are left blank

We analyzed the effect of the serum trace elements concentration on the ΔCT of each miRNA using multiple linear regression models, corrected for maternal age and pre-pregnancy BMI (Figure 3 Panels A-C). A significant positive association was found between increasing concentrations of Se and Zn and miR-223 ΔCT level (β_Se_=1.56, 95% CI_Se_=0.09–3.02; β_Zn_=1.67, 95% CI_Zn_=0.33–3.03). Conversely, an inverse association emerged between increasing Hg level and miR30-b ΔCT (β_Hg_=-0.66, 95% CI_Hg_=-1.27– -0.04). No specific association was observed between Let-7a expression and contaminants concentration.

**Figure 3.**
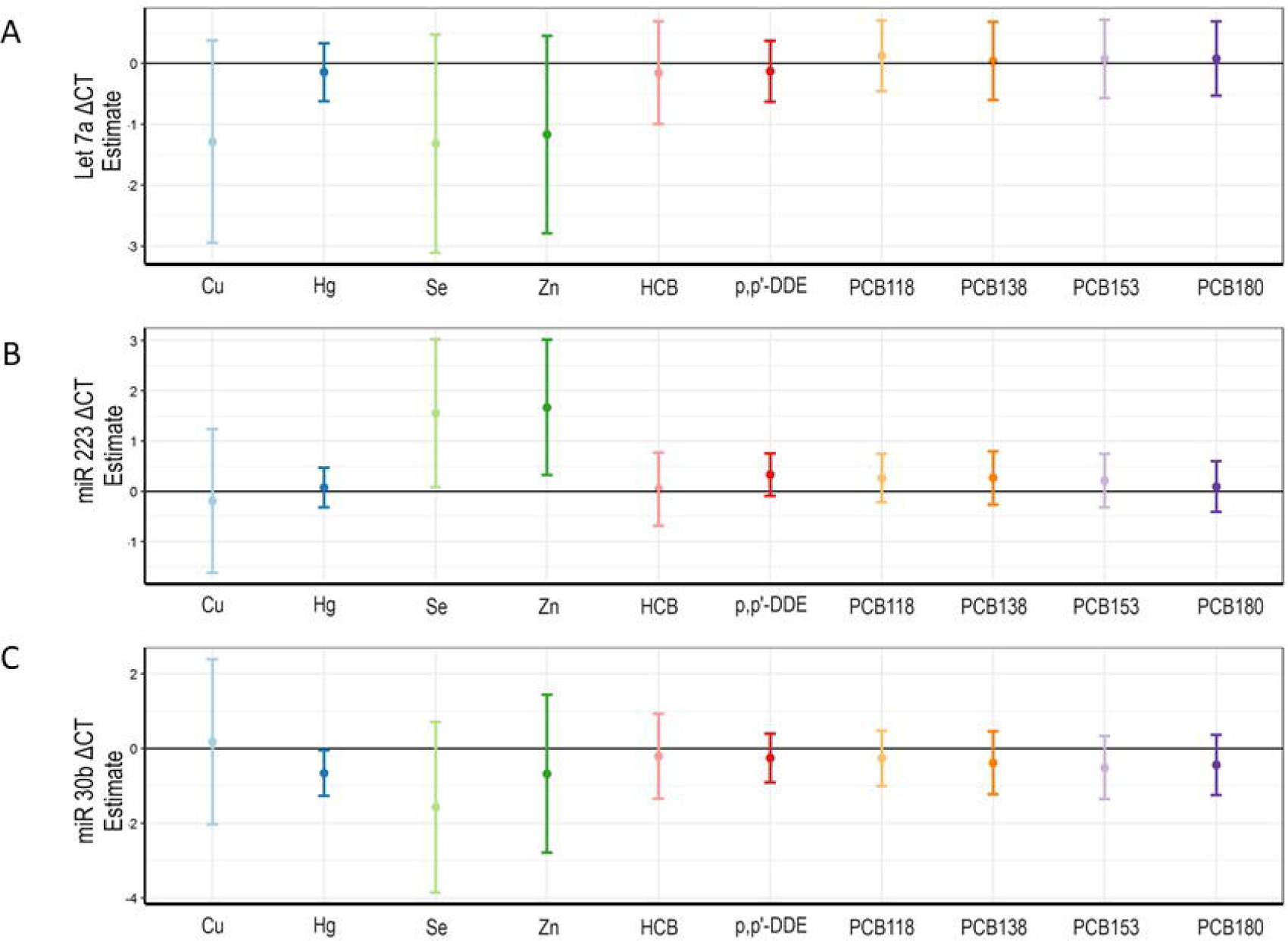
Linear regression models. Panels A-C show linear models for Let-7a, miR-223 and miR-30b, respectively. Each model was adjusted for maternal age and pre-pregnancy BMI.

To evaluate the association between tertiles of each contaminant and miRNA expression, multiple linear regression models were computed. As shown in Figure 4 Panel A, we found a significant Let-7a ΔCT reduction (β =-1.82; p=0.0009) in the comparison between the medium and low tertiles of Se. MiR-223 ΔCT showed a significant positive association with the high tertiles of Zn and p,p’-DDE (β_Zn_=1.02; p=0.024 and β_DDE_ =0.93 p= 0.050 -Figure 4, Panel B). As concerns miR-30b, we found a significant reduction of ΔCT in correspondence with the high tertile of Hg (β =-1.80; p=0.021) (Figure 4 panel C).

**Figure 4.**
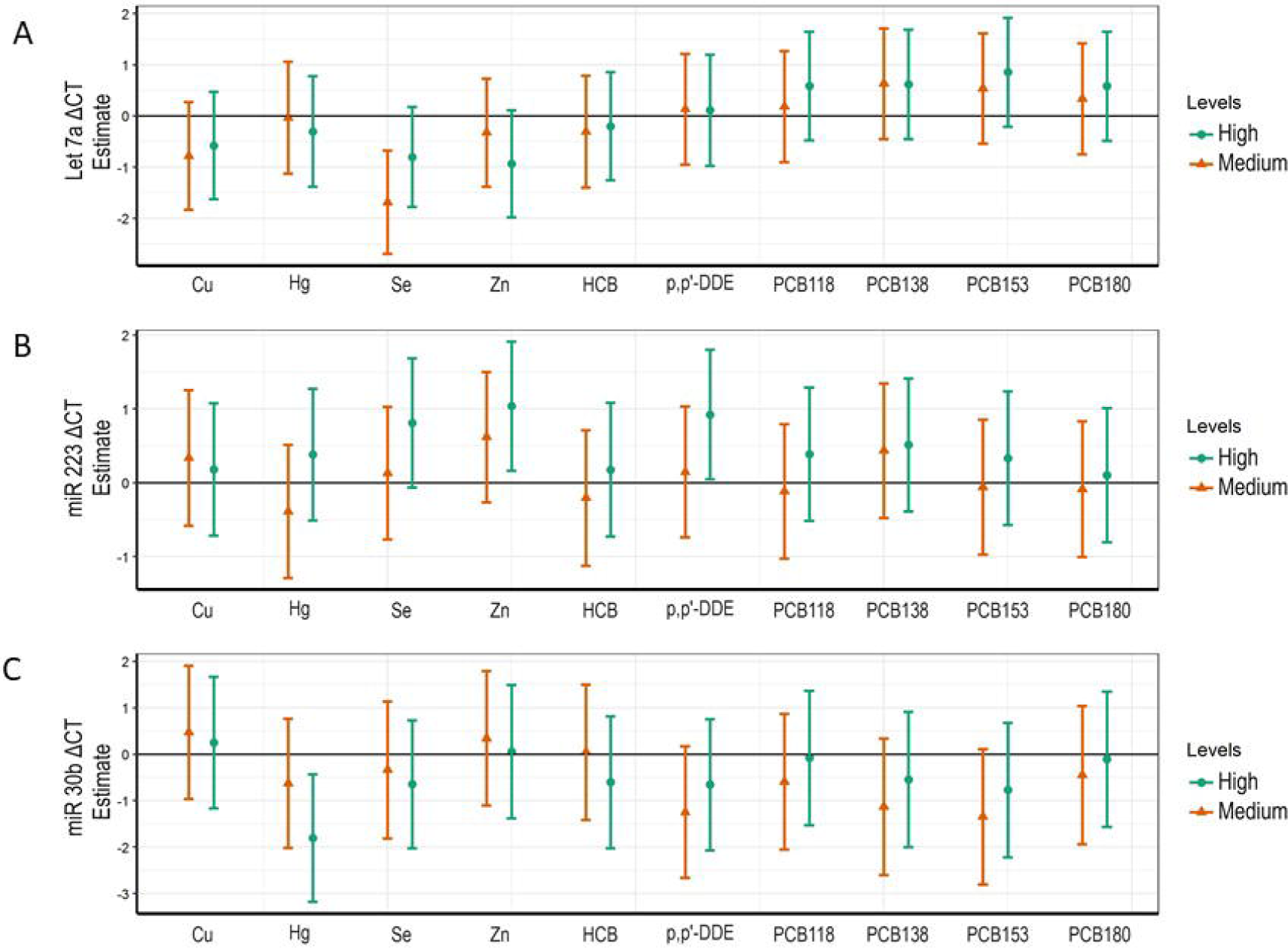
Multiple linear regression models for Let-7a, miR-223 and miR-30b ΔCTs. Panels A-C show linear models for Let-7a, miR-223 and miR-30b, respectively. Each exposure was categorized into tertile, lower tertile was used as reference category. Each model was adjusted for maternal age and pre-pregnancy BMI.

Furthermore, the expression levels for the selected miRNAs were also reported in fold change using the 2^-ΔΔCT^ method, showing a 3.2-fold increase in the expression of Let-7a in the medium vs low tertiles of Se. For miR-30b, we observed a 3.5-fold increase in the high vs low tertiles for Hg, a 2.4-fold increase in the medium vs low tertiles of p,p’-DDE, a 2.2 fold increase for PCB138 and a 2.5 fold increase for PCB 153. Conversely, we observed a 0.5 fold reduced expression of miR-223 in the high vs low tertiles for Zn and p,p’-DDE concentrations, respectively. Table 3 reports the miRNA expression values, in fold change, obtained from all toxicants and trace element concentration tertiles.

**Table 3.** MicroRNAs fold increase: green and red boxes show increased and decreased levels of expression, respectively.

|  |  | Let-7a | miR-223 | miR-30b |
| --- | --- | --- | --- | --- |
|  |  | Fold Change | Fold Change | Fold Change |
| Cu | Medium | 1.7 | 0.8 | 0.7 |
|  | High | 1.5 | 0.9 | 0.8 |
| Hg | Medium | 1.0 | 1.3 | 1.5 |
|  | High | 1.2 | 0.8 | 3.5 |
| Se | Medium | 3.2 | 0.9 | 1.3 |
|  | High | 1.7 | 0.6 | 1.6 |
| Zn | Medium | 1.3 | 0.7 | 0.8 |
|  | High | 1.9 | 0.5 | 1.0 |
| HCB | Medium | 1.2 | 1.2 | 1.0 |
|  | High | 1.1 | 0.9 | 1.5 |
| p,p'-DDE | Medium | 0.9 | 0.9 | 2.4 |
|  | High | 0.9 | 0.5 | 1.6 |
| PCB118 | Medium | 0.9 | 1.1 | 1.5 |
|  | High | 0.7 | 0.8 | 1.1 |
| PCB138 | Medium | 0.6 | 0.7 | 2.2 |
|  | High | 0.7 | 0.7 | 1.5 |
| PCB153 | Medium | 0.7 | 1.0 | 2.5 |
|  | High | 0.6 | 0.8 | 1.7 |
| PCB180 | Medium | 0.8 | 1.1 | 1.4 |
|  | High | 0.7 | 0.9 | 1.1 |

### Multipollutant analysis

According to the previous data reported in Figures 2-4, we decided to apply a multipollutant analysis to estimate the relative weights of the serum toxicants, allowing us to make inferences concerning the relative importance of exposures. We decided to build the WQS regression analysis on tertiles of distribution of each contaminant, according to low, medium, and high levels of exposure. The WQS regression showed a significant inverse association between Let-7a ΔCT and the WQS index (β= -1.005, p=0.034). The relative weights of Zn and Se predominated in the WQS index, followed by Cu, Hg, and HCB, indicating their importance in influencing Let-7a (Figure 5, Panel A) expression. As concerns miR-223, we found that the WQS index was significantly associated with an increase of ΔCT (β =1.369, p=0.005). Once again, Zn dominated the weights of the WQS index (Figure 5, Panel B). This was followed by the weight of p,p’-DDE and Se. Lastly, we also found a significant association between the WQS index and ΔCT reduction of miR-30b (β =-1.088, p=0.063), while the bad actors were represented by Hg, followed by Se (Figure 5, Panel C).

**Figure 5.**
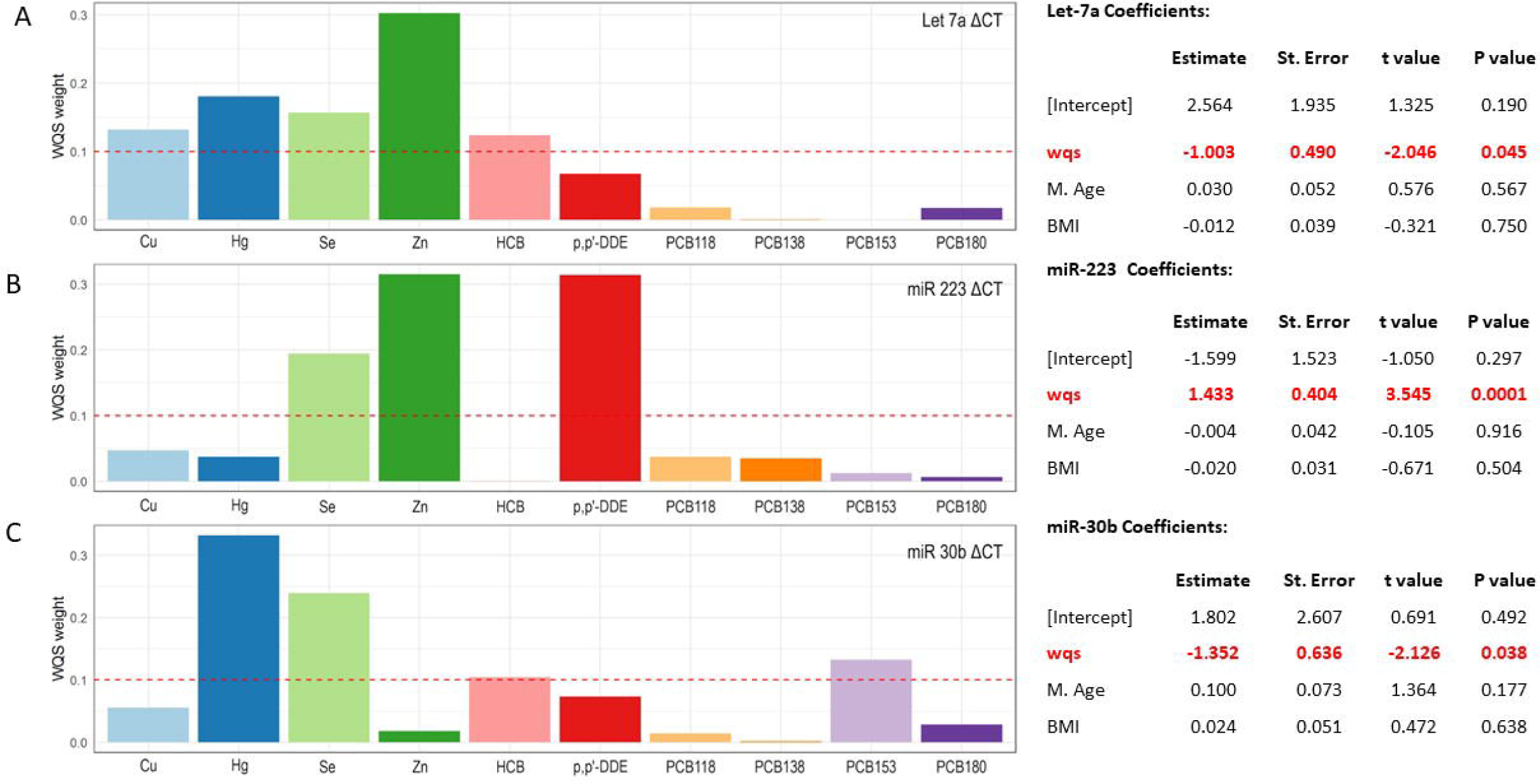
WQS regression analysis. Association between concentrations of biomonitored substances and Let-7a (Panel A), miR-223 (Panel B) and miR-30b (Panel C) ΔCTs based on weighted quantile sum (WQS) regression analysis.

## DISCUSSION

Humans are regularly exposed to a large number of environmental chemicals that have potentially toxic effects on health. Exposure to environmental agents may lead to epigenetic reprogramming, including changes in non-coding RNAs signatures, which can contribute to the induction of developmental changes or lead to disease progression and/or pathophysiology state (Miguel et al. 2020). Until now, most studies have focused on chemical exposures including cadmium (Brooks and Fry 2017), phthalates (LaRocca et al. 2016), arsenic, and endocrine disrupting chemicals (Sood et al. 2017) in association with placental miRNAs. Indeed, miRNAs are considered guardians of cellular homeostasis, regulating several processes including stress responses. Environmental pollutants could interact with the miRNA biosynthesis machinery, thereby directly inducing changes in miRNA expression levels (Izzotti and Pulliero 2014; Ligorio et al. 2011). In particular, miR-223 is widely involved in the regulation of several cellular processes such as cancer, acute lung injury, rheumatoid arthritis, autoimmune and inflammatory diseases (Haneklaus et al. 2013; Mancuso et al. 2019; Neudecker et al. 2017). MiR⍰30b, a member of the miR⍰30 family, has been implicated in the pathogenesis of multiple diseases, including various types of cancer, cardiovascular, renal and neurological disorders (Zhang et al. 2021). The Let-7 family of miRNAs has emerged as a central regulator of systemic energy homeostasis and it displays remarkable plasticity in metabolic responses to nutrients availability and physiological activities (Jiang 2019).

Following this line of evidence, we evaluated the maternal serum levels of relevant micro-RNAs such as Let-7a, miR-223 and miR-30b by Real Time PCR. First of all, we analyzed the level of expression of the selected microRNAs within the two groups of mothers (NPCS vs LRA) showing a non-significant association between the level of expression of the single miRNAs and the geographical localization participants’ dwellings (Table 1S and Figure 1S). Furthermore, a Pearson’s correlation matrix between the level of expression of the three miRNAs and the concentration of maternal serum contaminants allowed us to investigate the dependence between multiple variables in the selected mothers from the NEHO cohort. The result reported within Figure 2 shows the pairwise correlation coefficients between maternal serum biomonitoring data and the miRNAs with particular references to Hg, Se and Zn suggesting a correlation between the different toxicants and the expression level of the selected microRNAs. Starting from these observations, we analyzed the cohort in separate linear regression models for Let-7a, miR-223 and miR-30b adjusted for maternal age and pre-pregnancy BMI. In this setting, we observed a significant association between the level of expression of miR-223 and the serum concentration of Se and Zn and an inverse association between the expression of miR-30b and the serum concentration of Hg (Figure 3). Then, we applied separate multiple linear regression models to the groups of contaminants and miRNAs exploring the specific effects of each pollutant, categorized into tertiles according to low, medium and high levels of exposure. In each model, the lower tertile was used as reference. According to this strategy, we observed that Let-7a showed a significant reverse association with the medium level of Se while the higher tertile of Zn and p,p’-DDE showed significant positive associations with the miR-223 ΔCT. Moreover, the higher tertile of Hg presented an inverse association with miR-30b (see Figure 4 panels A-C for details).

These findings were further confirmed as shown within Table 3, where the correlation between miRNAs response and the Low, Medium and High tertiles of each pollutant were reported as fold changes in terms of miRNAs expression. In particular, a significant positive increase of Let-7a expression is also evident in the medium vs low tertiles of Se and in the miR-30b in the high vs low tertiles for Hg and in the medium vs low tertiles of p,p’-DDE, PCB138 and PCB 153. Differently, a significant reduced expression emerges for the miR-223 in the high vs low tertiles for Zn and p,p’-DDE concentration (see Table 3 for details).

However, this does not exclude indirect interactions between independent stressors and synergistic impact on health even at concentrations lower than their individual recognized toxicity thresholds. Some chemicals at relatively lower toxicity doses may likewise be insidious (Barouki et al. 2012), mainly during critical developmental periods of humans, where long-lasting exposures may induce subtle epigenetic alterations leading to permanent changes that can possibly program health trajectories throughout the rest of lifespan. Thus, we explored the effect of the complex environmental mixtures of contaminants using the Weighted Quantile Sum (WQS) statistical regression model (Lee et al. 2019; Tanner et al. 2020). This model constructs a weighted index estimating the mixed effect of all predictor variables on an outcome, and the WQS regression result indicates the respective importance of individual chemicals related to specific outcomes, thus identifying driving elements also called “bad actors”. The contribution of each individual predictor to the overall index effect may then be assessed by the relative strength of the weights the model assigns to each variable. By means of mixture weight, we create a weighted index as a measure of a mixture effect able to identify the relative weight of the different investigated substances on the level of expression of circulating Let-7a, miR-223 and miR-30b miRNAs. The WQS regression analysis showed a significant inverse association between Let-7a ΔCT and the relative weights of Zn, Se, Cu, Hg and HCB indicating their importance in influencing Let-7a expression (Figure 5 panel A). As concerns miR-223, we found that the WQS index was significantly associated with the increase of ΔCT where Zn and p,p’-DDE dominated the weights of the WQS index followed by the Se (Figure 5 panel B). Lastly, we also found an association between the WQS index and ΔCT reduction of miR-30b in association manly to Hg and Se levels followed by the PCB153 (Figure 5 panel C). These preliminary results clearly suggest a real-life exposure to mixtures that women experienced in a specific window of their pregnancy. The ensemble of this statistical approaches demonstrates the role played by human exposure to mixtures of contaminants in the environment with respect to single chemical impact (Braun et al. 2016). Thus, although the measured chemicals in the NPCS and LRA mothers were generally lower than those reported in studies in highly contaminates areas (Bocca et al. 2010) (Table 2), evidence of exposure to the mixtures of contaminants appears statistically measurable in terms of microRNAs expression. Noteworthy, these effects appear from healthy mothers, with no previous report of preterm delivery or other pre-existent pathologies. This implies that significant epigenetic changes, modulating the expression of some microRNAs, represent a reliable clue for risk assessment of diseases in both mothers and newborns. This emerges as a decisive outcome considering the reduced number of pollutants measured in this research with respect +to the wider list of chemicals reported from Augusta area, with their potential synergistic combination and definitively suggests a critical role that biomonitoring of microRNAs could play in terms of hazard evaluation of environmental pollution on human health. These epigenetic modifications integrate, in a sort of synthetic responses, the effect of cocktails of pollutants in multi-environmental matrices on exposed individuals.

Understanding the molecular mechanisms by which a large set of different substances such as dietary factors and/or environmental chemicals can influence the expression of circulating microRNAs is an active area of research (Ross and Davis 2014). The reported results allowed the identification of the relative weight of several different substances which seems to suggest the importance of the redox dependent regulation for the maintenance of cell homeostasis and the role of essential elements and miRNAs in this process. In fact, during pregnancy a higher frequency of oxidative reactions have been reported (Bakacak et al. 2015) and the involvement of essential elements and of the Let-7a, miR-223 and miR-30b have also been described (Ciesielska et al. 2021; Kalinina et al. 2019). The mutual feedback regulation of ROS and miRNAs, i.e., the interplay between the ROS action on expression of miRNAs and control of the ROS level via miRNAs expression is beyond the aim of our study but a better comprehension of the hierarchy of redox-dependent regulatory systems for the cell functioning is certainly of paramount importance for future studies.

## Data Availability

All data produced in the present work are contained in the manuscript

## Figure Legend

**Figure 1s. EDA plots of serum miRNA** Δ**CTs**. Panels A-C display the distribution of the Let-7a; miR-30b, miR223 microRNAs, respectively. The histograms report the relative frequency distribution of all considered miRNAs. The box-plot, from left to right, shows, 5th, 25th, 50th (median), 75th and 95th percentile. Red dots and blue dots represent subjects from Local Reference areas and National Priority Contaminated Sites, respectively.

## Acknowledgments

This Study was funded by the International Centre of advanced Study in Environment, ecosystem and human Health (CISAS), a multidisciplinary project on environment/health relationships funded by the Italian Ministry of Education, Universities and Research (MIUR) and approved by the Interministerial Committee for Economic Planning (CIPE) – body of the Italian Government – with Resolution no. 105/2015 of December 23, 2015

## REFERENCE

Apicella C, Ruano CSM, Mehats C, Miralles F, Vaiman D. 2019. The role of epigenetics in placental development and the etiology of preeclampsia. International journal of molecular sciences 20.

Arck PC, Hecher K. 2013. Fetomaternal immune cross-talk and its consequences for maternal and offspring’s health. Nature medicine 19:548–556.

Bagnato E, Sproveri M, Barra M, Bitetto M, Bonsignore M, Calabrese S, et al. 2013. The sea-air exchange of mercury (hg) in the marine boundary layer of the augusta basin (southern italy): Concentrations and evasion flux. Chemosphere 93:2024–2032.

Bakacak M, Kilinc M, Serin S, Ercan O, Kostu B, Avci F, et al. 2015. Changes in copper, zinc, and malondialdehyde levels and superoxide dismutase activities in pre-eclamptic pregnancies. Medical science monitor : international medical journal of experimental and clinical research 21:2414–2420.

Barker DJ. 1993. Fetal origins of coronary heart disease. British heart journal 69:195–196.

Barouki R, Gluckman PD, Grandjean P, Hanson M, Heindel JJ. 2012. Developmental origins of non-communicable disease: Implications for research and public health. Environmental health : a global access science source 11:42.

Bellucci LG, Giuliani S, Romano S, Albertazzi S, Mugnai C, Frignani M. 2012. An integrated approach to the assessment of pollutant delivery chronologies to impacted areas: Hg in the augusta bay (italy). Environmental science & technology 46:2040–2046.

Bocca B, Mattei D, Pino A, Alimonti A. 2010. Italian network for human biomonitoring of metals: Preliminary results from two regions. Annali dell’Istituto superiore di sanita 46:259–265.

Bocca B, Ruggieri F, Pino A, Rovira J, Calamandrei G, Mirabella F, et al. 2020. Human biomonitoring to evaluate exposure to toxic and essential trace elements during pregnancy. Part b: Predictors of exposure. Environmental research 182:109108.

Boosani CS, Agrawal DK. 2016. Epigenetic regulation of innate immunity by micrornas. Antibodies 5.

Braun JM, Gennings C, Hauser R, Webster TF. 2016. What can epidemiological studies tell us about the impact of chemical mixtures on human health? Environmental health perspectives 124:A6–9.

Brooks SA, Fry RC. 2017. Cadmium inhibits placental trophoblast cell migration via mirna regulation of the transforming growth factor beta (tgf-beta) pathway. Food and chemical toxicology : an international journal published for the British Industrial Biological Research Association 109:721–726.

Carere M, Musmeci L, Comba P, Bianchi F, Lepore V, Pilozzi A. 2016. Study for the environmental health characterization of the contaminated sites of gela and priolo. Rapporti ISTISAN 2016.

Ciesielska S, Slezak-Prochazka I, Bil P, Rzeszowska-Wolny J. 2021. Micro rnas in regulation of cellular redox homeostasis. International journal of molecular sciences 22.

Davies S, Briand V, Accrombessi M, Fievet N, Le Bot B, Durand S, et al. 2021. Pre-conception serum ferritin concentrations are associated with metal concentrations in blood during pregnancy: A cohort study in benin. Environmental research 202:111629.

Deng Q, Lu C, Li Y, Sundell J, Dan N. 2016. Exposure to outdoor air pollution during trimesters of pregnancy and childhood asthma, allergic rhinitis, and eczema. Environmental research 150:119–127.

Dennis KK, Marder E, Balshaw DM, Cui Y, Lynes MA, Patti GJ, et al. 2017. Biomonitoring in the era of the exposome. Environmental health perspectives 125:502–510.

Di Bella C, Traina A, Giosue C, Carpintieri D, Lo Dico GM, Bellante A, et al. 2020. Heavy metals and pahs in meat, milk, and seafood from augusta area (southern italy): Contamination levels, dietary intake, and human exposure assessment. Frontiers in public health 8:273.

Drago G, Ruggieri S, Bianchi F, Sampino S, Cibella F. 2020. Birth cohorts in highly contaminated sites: A tool for monitoring the relationships between environmental pollutants and children’s health. Frontiers in public health 8:125.

Ekino S, Susa M, Ninomiya T, Imamura K, Kitamura T. 2007. Minamata disease revisited: An update on the acute and chronic manifestations of methyl mercury poisoning. Journal of the neurological sciences 262:131–144.

Feo ML, Bagnati R, Passoni A, Riva F, Salvagio Manta D, Sprovieri M, et al. 2020. Pharmaceuticals and other contaminants in waters and sediments from augusta bay (southern italy). The Science of the total environment 739:139827.

Haneklaus M, Gerlic M, O’Neill LA, Masters SL. 2013. Mir-223: Infection, inflammation and cancer. Journal of internal medicine 274:215–226.

Izzotti A, Pulliero A. 2014. The effects of environmental chemical carcinogens on the microrna machinery. International journal of hygiene and environmental health 217:601–627.

Jiang S. 2019. A regulator of metabolic reprogramming: Microrna let-7. Translational oncology 12:1005–1013.

Junien C, Panchenko P, Pirola L, Amarger V, Kaeffer B, Parnet P, et al. 2016. [the new paradigm of the developmental origin of health and diseases (dohad)--epigenetics and environment: Evidence and missing links]. Med Sci (Paris) 32:27–34.

Kalinina EV, Ivanova-Radkevich VI, Chernov NN. 2019. Role of micrornas in the regulation of redox-dependent processes. Biochemistry Biokhimiia 84:1233–1246.

Lamichhane DK, Leem JH, Lee JY, Kim HC. 2015. A meta-analysis of exposure to particulate matter and adverse birth outcomes. Environmental health and toxicology 30:e2015011.

LaRocca J, Binder AM, McElrath TF, Michels KB. 2016. First-trimester urine concentrations of phthalate metabolites and phenols and placenta mirna expression in a cohort of u.S. Women. Environmental health perspectives 124:380–387.

Lee M, Rahbar MH, Samms-Vaughan M, Bressler J, Bach MA, Hessabi M, et al. 2019. A generalized weighted quantile sum approach for analyzing correlated data in the presence of interactions. Biometrical journal Biometrische Zeitschrift 61:934–954.

Ligorio M, Izzotti A, Pulliero A, Arrigo P. 2011. Mutagens interfere with microrna maturation by inhibiting dicer. An in silico biology analysis. Mutation research 717:116–128.

Longo V, Longo A, Di Sano C, Cigna D, Cibella F, Di Felice G, et al. 2019. In vitro exposure to 2,2’,4,4’-tetrabromodiphenyl ether (pbde-47) impairs innate inflammatory response. Chemosphere 219:845–854.

Longo V, Longo A, Adamo G, Fiannaca A, Picciotto S, La Paglia L, et al. 2021. 2,2’4,4’-tetrabromodiphenyl ether (pbde-47) modulates the intracellular mirna profile, sev biogenesis and their mirna cargo exacerbating the lps-induced pro-inflammatory response in thp-1 macrophages. Frontiers in immunology 12:664534.

Louro H, Heinala M, Bessems J, Buekers J, Vermeire T, Woutersen M, et al. 2019. Human biomonitoring in health risk assessment in europe: Current practices and recommendations for the future. International journal of hygiene and environmental health 222:727–737.

Mancuso R, Agostini S, Hernis A, Zanzottera M, Bianchi A, Clerici M. 2019. Circulatory mir-223-3p discriminates between parkinson’s and alzheimer’s patients. Scientific reports 9:9393.

Miguel V, Lamas S, Espinosa-Diez C. 2020. Role of non-coding-rnas in response to environmental stressors and consequences on human health. Redox biology 37:101580.

Montalbano AM, Albano GD, Anzalone G, Moscato M, Gagliardo R, Di Sano C, et al. 2020. Cytotoxic and genotoxic effects of the flame retardants (pbde-47, pbde-99 and pbde-209) in human bronchial epithelial cells. Chemosphere 245:125600.

Mudu P, Terracini B, Martuzzi M. 2014. Human health in areas with industrial contamination. World Health Organization.

Neudecker V, Haneklaus M, Jensen O, Khailova L, Masterson JC, Tye H, et al. 2017. Myeloid-derived mir-223 regulates intestinal inflammation via repression of the nlrp3 inflammasome. The Journal of experimental medicine 214:1737–1752.

Rahman SM, Kippler M, Ahmed S, Palm B, El Arifeen S, Vahter M. 2015. Manganese exposure through drinking water during pregnancy and size at birth: A prospective cohort study. Reproductive toxicology 53:68–74.

Rappaport SM, Barupal DK, Wishart D, Vineis P, Scalbert A. 2014. The blood exposome and its role in discovering causes of disease. Environmental health perspectives 122:769–774.

Rappaport SM. 2016. Genetic factors are not the major causes of chronic diseases. PloS one 11:e0154387.

Ross SA, Davis CD. 2014. The emerging role of micrornas and nutrition in modulating health and disease. Annual review of nutrition 34:305–336.

Ruggieri S, Drago G, Colombo P, Alesci A, Augello P, Bisbano A, et al. 2019. Three contaminated sites in southern italy. The neonatal environment and health outcomes cohort: Protocol for a longitudinal birth cohort study. BMJ open 9:e029471.

Ruggieri S, Maltese S, Drago G, Cibella F, Panunzi S. 2021. The neonatal environment and health outcomes (neho) birth cohort study: Behavioral and socioeconomic characteristics and drop-out rate from a longitudinal birth cohort in three industrially contaminated sites in southern italy. International journal of environmental research and public health 18.

Salvagio Manta D, Bonsignore M, Oliveri E, Barra M, Tranchida G, Giaramita L, et al. 2016. Fluxes and the mass balance of mercury in augusta bay (sicily, southern italy). Estuarine, Coastal and Shelf Science 181: 134–143.

Sood S, Shekhar S, Santosh W. 2017. Dimorphic placental stress: A repercussion of interaction between endocrine disrupting chemicals (edcs) and fetal sex. Medical hypotheses 99:73–75.

Sprovieri M, Oliveri E, Di Leonardo R, Romano E, Ausili A, Gabellini M, et al. 2011. The key role played by the augusta basin (southern italy) in the mercury contamination of the mediterranean sea. Journal of environmental monitoring : JEM 13:1753–1760.

Stolevik SB, Nygaard UC, Namork E, Haugen M, Meltzer HM, Alexander J, et al. 2013. Prenatal exposure to polychlorinated biphenyls and dioxins from the maternal diet may be associated with immunosuppressive effects that persist into early childhood. Food and chemical toxicology : an international journal published for the British Industrial Biological Research Association 51:165–172.

Tanner E, Lee A, Colicino E. 2020. Environmental mixtures and children’s health: Identifying appropriate statistical approaches. Current opinion in pediatrics 32:315–320.

Traina A, Ausili A, Bonsignore M, Fattorini D, Gherardi S, Gorbi S, et al. 2021. Organochlorines and polycyclic aromatic hydrocarbons as fingerprint of exposure pathways from marine sediments to biota. Marine pollution bulletin 170:112676.

Wild CP. 2005. Complementing the genome with an “exposome”: The outstanding challenge of environmental exposure measurement in molecular epidemiology. Cancer Epidemiol Biomarkers Prev 14:1847–1850.

Wu Y, Li Q, Zhang R, Dai X, Chen W, Xing D. 2021. Circulating micrornas: Biomarkers of disease. Clinica chimica acta; international journal of clinical chemistry 516:46–54.

Zhang Q, Liu S, Zhang J, Ma X, Dong M, Sun B, et al. 2021. Roles and regulatory mechanisms of mir-30b in cancer, cardiovascular disease, and metabolic disorders (review). Experimental and therapeutic medicine 21:44.

Zona A, Iavarone I, Buzzoni C, Conti S, Santoro M, Fazzo L, et al. 2019. [sentieri: Epidemiological study of residents in national priority contaminated sites. Fifth report]. Epidemiologia e prevenzione 43:1–208.

