## Supplementary figures and images for "Environmental pollutants and essential elements as regulators of miR-30b, miR-223 and Let-7a microRNAs expression in maternal sera from the NEHO cohort"

### supplementary Figure 1s

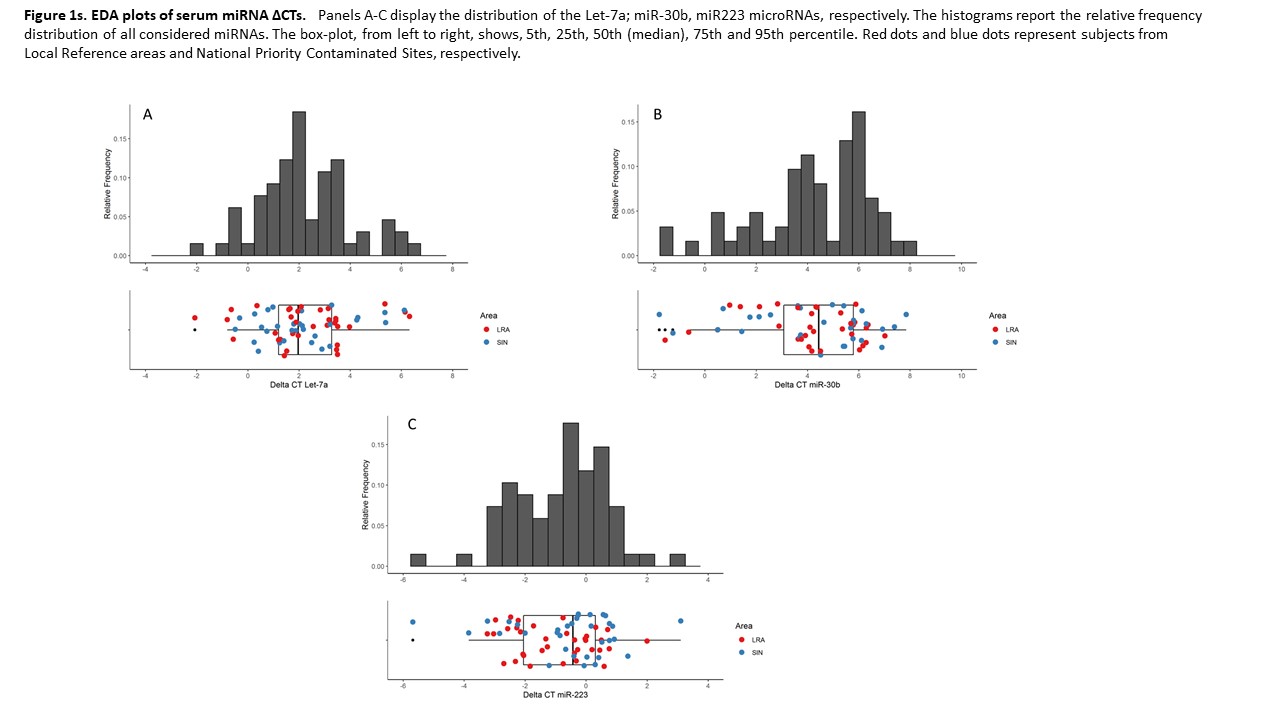

### supplementary table 1s

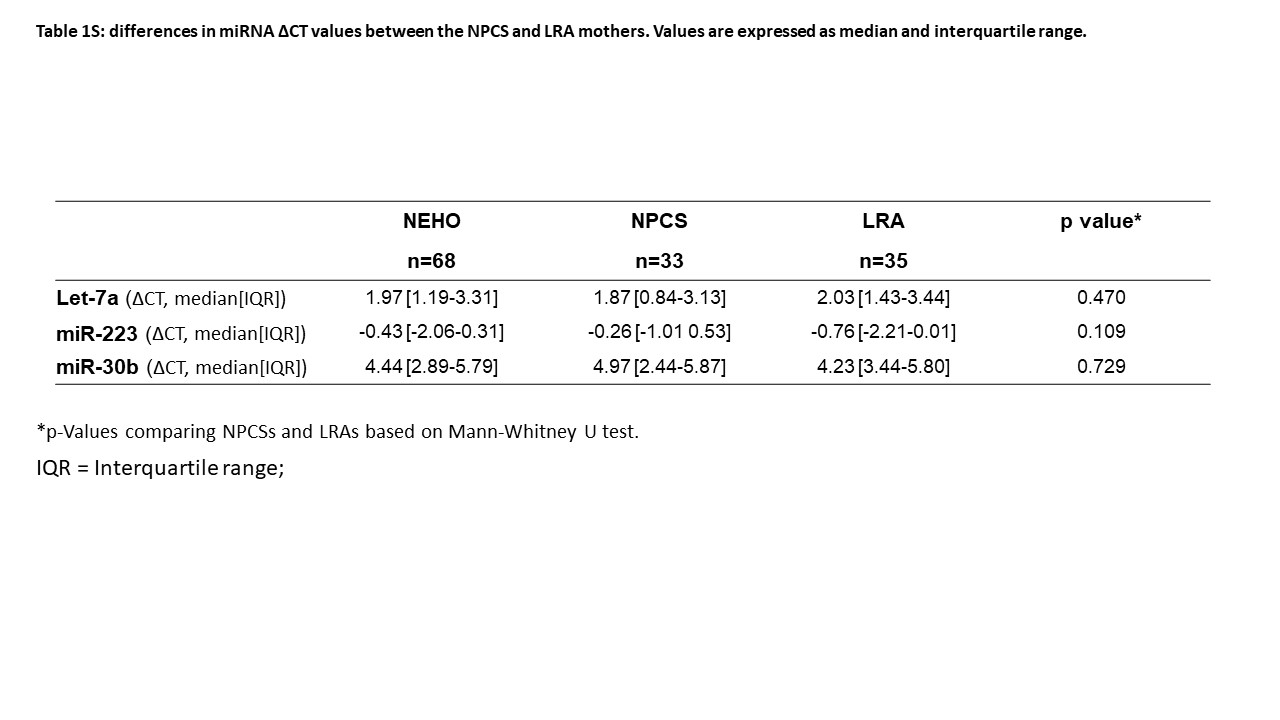
